# Mathematical and Statistical models as tools for the control of mosquito-borne diseases: The experience of Costa Rica

**DOI:** 10.1101/2021.06.24.21259470

**Authors:** Paola Vásquez, Fabio Sanchez, Luis A. Barboza, Yury E. García, Juan G. Calvo, Shu-Wei Chou, Gustavo Mery

## Abstract

The modeling of infectious diseases provides valuable input for developing mitigating strategies and implementing public health interventions. This article highlights results and current research conducted in Costa Rica using mathematical, statistical, and computational tools to analyze transmission dynamics and provide quantitative information for developing mosquito-borne disease prevention/control methods and resource allocation planning.

## 1 Introduction

Currently, despite strong control efforts and the increased knowledge acquired throughout history, timely and effective management of mosquito-borne pathogens continue to elude public health authorities worldwide. The past decades have been especially challenging as increased urbanization, population mobility, deforestation, climate change, insecticide resistance, and deficient vector control programs have created viable conditions for both pathogens and vectors to emerge into new areas and to reemerge in regions from which previously successful vector control campaigns were implemented.

This scenario has led public health officials worldwide to highlight the need to explore innovative and cost-effective tools to strengthen the prevention and mitigation programs in affected countries. However, the complexity involved in the transmission dynamics, coupled with newly recognized pathogenic mechanisms and modes of transmission, urgently demands a more interdisciplinary approach to reach this goal [1]. In this effort, policymakers have recognized the use of mathematical, computational, and statistical modeling techniques as essential and plausible tools for vector control [2].

In essence, these models provide a simplified representation of a complex system, which involves various underlying factors, interactions, heterogeneity, random variations, and fluctuations. They allow us to either improve the understanding of how a specific pathogen spreads, test a diversity of transmission scenarios and mitigation actions, evaluate the impact of control strategies in disease incidence, or provide projections of possible transmission tendencies to better allocate available resources [3]. For their development, multiple modeling techniques and data have been used [6], including climate, environmental, entomological, demographic, and socioeconomic variables. The results obtained have shown the potential of such models to emphasize critical factors for public health interventions and to guide public policy, thus helping administer human and economic resources more efficiently for the fight against mosquito-borne pathogens [2].

However, every model has limitations intrinsic to the model itself, to the user experience, or the availability of information. Therefore, several challenges arise when attempting to use them for public health purposes. Nevertheless, the increased knowledge in mosquito population biology, advances in technology, access to the enormous amount of real-time epidemiological, demographic, socioeconomic, and climate data have opened up a new array of possibilities for these models to reinvigorate disease control [1].

In Costa Rica, vector-borne diseases have been a burden for public health authorities for decades. Since the introduction of the dengue virus in 1993 up to 2020 more than 392,000 dengue cases have been reported by the Ministry of Health [4]. In 2014, the Ministry of Health reported the first cases of chikungunya; by February 2016, the first indigenous Zika cases appeared on the Pacific coast, and since 2016 the local transmission of malaria has slowly resumed its upwards trend [4]. Therefore, the introduction and development of new cost-effective methods and tools are at the forefront of public health priorities in the country.

Here, we reviewed four modelling techniques used in Costa Rica that range from the classic compartmental epidemic models and Approximate Bayesian Computation (ABC) to more complex computational machine learning tools and network models with multi-level systems. For the ABC approach and machine learning methods, we provide the results from research conducted at the Research Center of Pure and Applied Mathematics (CIMPA) of the University of Costa Rica, and briefly mention the results from the other methods. The models were used to analyze the dynamics involved in the Chikungunya and Zika virus transmission and retrospectively predict the relative risk of acquiring dengue in five climatically diverse municipalities during 2017. We also include a summary of a network model currently used for projecting Covid-19 transmission in Costa Rica and its potential use to study vector-borne diseases [5].

## 2 Materials and Methods

The four methodologies presented in the article were developed using publicly available data from the Ministry of Health and other public institutions in Costa Rica. In order to fit the models with the available information, different statistical methods were used to infer the underlying parameters and state variables of each of the models described below.

### 2.1 Classical Methods

In studying the transmission dynamics of an infectious disease in a susceptible population, the use of a compartmental approach is considered the classical method of epidemic modelling. This approach, is based in dividing the population into different compartments according to their epidemiological status. The compartments range from the most basic division that includes Susceptible (S), Infectious (I), and Recovered (R) individuals to those with additional partitions and more complex interactions between classes. The dynamics of the disease are then characterized by either a system of ordinary differential equations (deterministic approach) or a stochastic process that calculates the periods and transitions between classes providing as a result tendencies of disease incidence in a given population.

In 2014, with the introduction of the Chikungunya virus into national territory, Costa Rican health authorities faced the circulation of two pathogens with similar clinical manifestations, temporal and spatial distribution, as the dengue virus has been endemic since 1993. Under this scenario, the use of a single-outbreak deterministic model, allowed to study and compare the trend of dengue and chikungunya reported cases in Costa Rica during the 2015-2016 outbreak [7].

For the analyses data regarding the number of cases was obtained from the Ministry of Health. The estimated parameters were the transmission rate, the diagnosis rate, the average vector infectious period, and the initial value of the susceptible population. The primary assumption was that the observed number of cases at week *k* follows a Poisson distribution whose rate is determined as a function of the expected number of cases within the *k*-th week. The authors used a least-squares procedure based on the normalized differences between observed and expected weekly cases to fit the whole set of parameters and initial populations.

### 2.2 Approximate Bayesian Computation

The Approximate Bayesian computation (ABC) is a type of computational technique that has its roots in the Bayesian statistics approach of data analysis and parameter estimation. Under this technique, the parameters of the model are not chosen deterministically, but sampled from a probability distribution, thus having a wide range of applications in the analysis of complex problems arising in public health, such as the emergence of the zika virus in the Americas in 2015.

In Cost Rica, by January 2016, the first cases of Zika were detected, adding yet another pathogen transmitted by the *Aedes* mosquito, with the added complexity of new ways of transmission and the congenital disabilities and pregnancy complications related to the virus.

With the combination of a Bayesian statistical approach and a single-outbreak mathematical model with sexual transmission, authors in [8] described the overall dynamics of Zika during 2016-2017 outbreak in Costa Rica. The estimated parameters were the mosquito’s average lifespan, the mosquito biting rate, and the per-capita diagnosis rate. From a Bayesian point of view, the parameters were treated as random variables whose posterior distribution was determined once we adopt a prior distribution for each of them. The authors overcame the limitation of the usual Bayesian estimation technique, which requires a well-defined likelihood function to express the posterior distribution of the parameters, by using an Approximate Bayesian computation approach with a rejection sampling scheme.

### 2.3 Statistical Learning

A different question of interest is projecting disease incidence based on past information from various sources. To address this question, the authors in [9] used nine years of dengue cases (2007-2016) provided by the Ministry of Health, as well as data of temperature, humidity, and rainfall provided by the National Meteorological Institute from five cantons across the country to feed two different machine learning approaches: Generalized Additive Model and Random Forest in an effort to retrospectively predict the relative risk of dengue in 2017.

Both methods belong to a more extensive set of Machine Learning techniques. This branch of statistical learning, represent a comprehensive set of algorithms that allows solving different tasks by learning from past transmission trends and dynamics. For example, prediction of dependent variables based on covariate behavior (Supervised Learning models) and pattern recognition over spaces of observations or variables (Non-supervised Learning models). Algorithms under this approach process data without instructions. explicit external (and therefore potentially biased) provided by the researcher.

In Costa Rica, these techniques are still in their early stages. However, the study conducted with dengue and climate variables in five municipalities of the country has shown promising results [9].

### 2.4 Network model

Complex systems can be modeled via network models. This type of modeling allows for complex heterogeneity, including the attribution of different characteristics for each node (individual) and its interactions among contacts (connected nodes in the graph). It can also incorporate multiple layers (distinct social groups) with different parameters per layer. With the emergence of the SARS-CoV-2 virus, and its the rapid propagation around the world, this type of models are a flexible tool to develop a wide range of transmission scenarios and project tendencies. In Costa Rica, a network model that uses the number of cases reported by the Ministry of Health and demographic information from the National Institute of Statistics and Census is used study various scenarios to project Covid-19 transmission in Costa Rica [5]. For vector-borne diseases, this framework will include a vector layer, which allows keeping track of its population and the interactions among them and between humans, per municipality. It also contains an underlying SIR-type epidemiological model to mimic the evolution of the disease, allowing a generalization of these types of models with less restrictive assumptions and more complex dynamics.

## 3 Results

By using classical methods to fit the tendencies of dengue and chikungunya reported cases in 2015, the model predictions were able to detect, during the months of May and June, a lower number of dengue cases than those reported by the Ministry of Health during the same period.

The rate at which individuals that were in the undiagnosed compartment became diagnosed as a dengue case was also calculated. Point estimates of key model parameters showed that individuals were being diagnosed at a faster rate for the dengue virus. Both data anomalies suggest that dengue cases were over reported in the 2015–2016 outbreak [7].

On the other hand, the Approximate Bayesian Computation approach allowed to demonstrate that people most at risk of becoming infected by the Zika virus were those who spent most of their time inside their homes. As shown in Figure 1, the number of secondary infections caused by an infected individual during his/her infectious period, ℛ_0_, dropped when the time spent inside their homes decreased.

**Figure 1:**
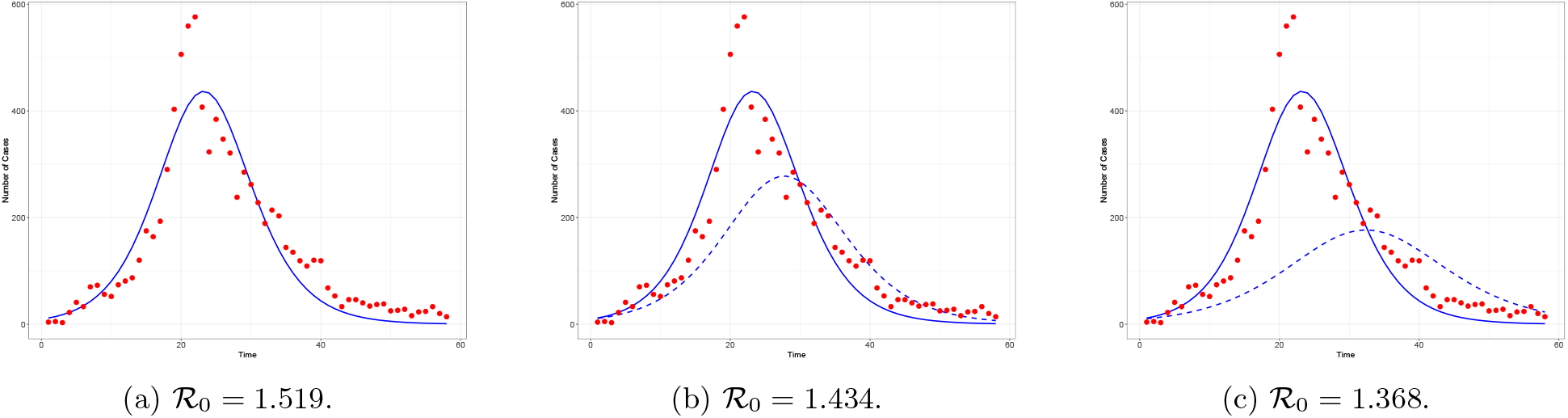
Number of reported Zika cases (red dots) from the 2016-2017 ZIKV outbreak in Costa Rica and model solutions. The solid blue line represents the best fit and the dotted line represents a lower percentage of individuals that stay indoors, therefore reducing the number of secondary infections [8].

In the efforts to predict the relative risk of dengue in Costa Rica, the machine learning algorithms showed an adequate performance in estimating the relative risk of dengue during 2017, while using the nine year period from 2007-2016 as training periods.

Figure 2 shows the results of the two different machine learning models. The dotted and solid lines correspond to the predicted relative risk of each study area over the testing period.

**Figure 2:**
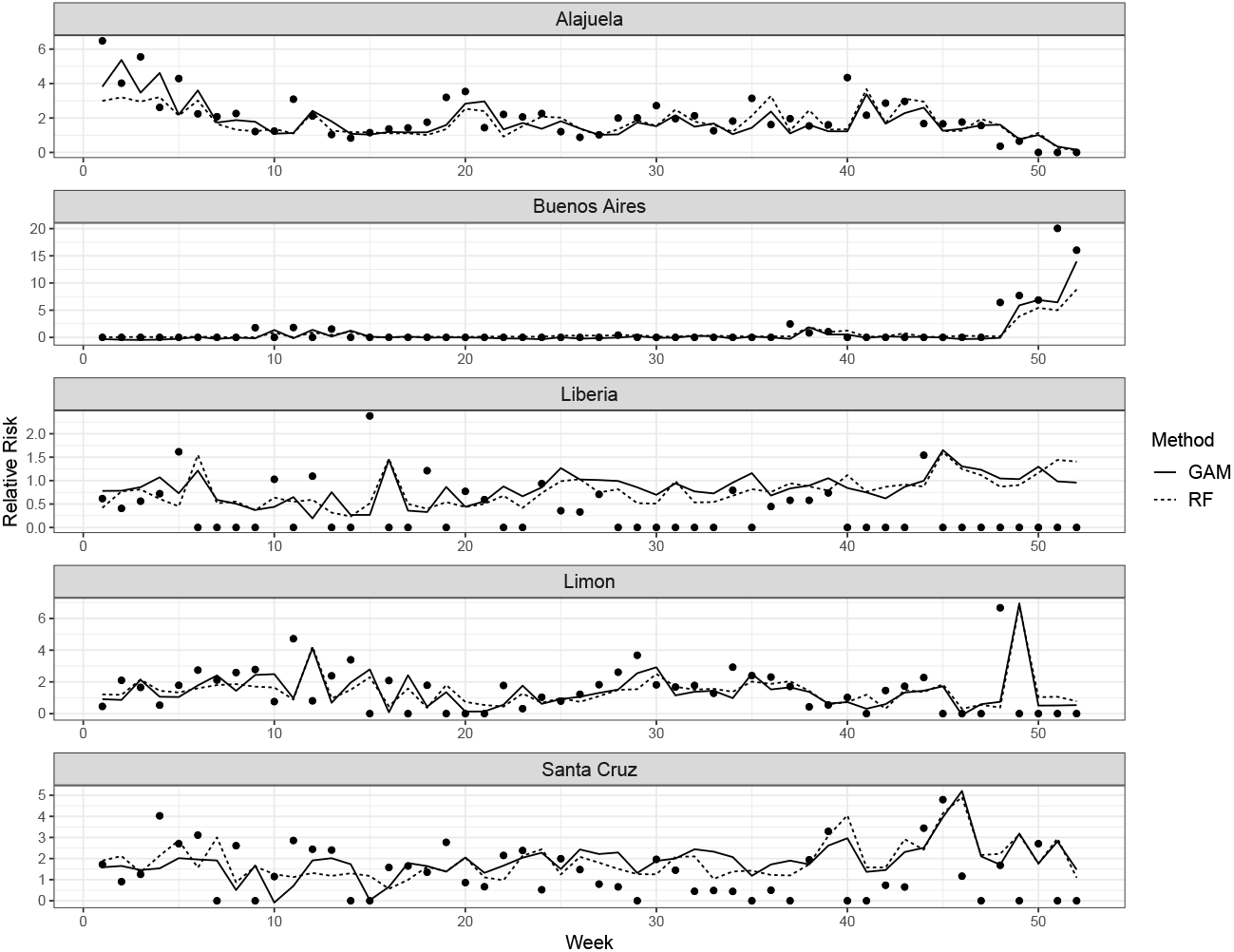
Model comparison over the prediction period [9]. Lines: predicted relative risk. Points: observed relative risk.

As seen in Figure 2, the models were able to predict the relative risk of dengue in the cantons of Limón, Buenos Aires, and Alajuela. These models also successfully predicted the general decreasing and increasing tendency of cases within weeks of precision. With the cantons on the North Pacific coast, Santa Cruz, and Liberia, the models overestimated the reported relative risk in most weeks. However, they could still predict weeks with an increasing or decreasing incidence precisely [9].

With the computer algorithms involved in the development of multilayer network models, the available epidemiological and demographic information has allowed to project the tendency of new SARS-CoV-2 cases and hospitalizations under a range of scenarios. In this context, and as showed in [5], the network model accurately projected that the wave caused by the introduction of more transmissible variants such as B.1.617.2 (Delta), coupled with different levels of vaccine hesitance, was going to be similar to the infection wave Costa Rica faced in May 2021 [5].

## 4 Discussion

In an increasingly interconnected and ever-changing world, public health systems of countries must be prepared to detect and respond quickly and effectively to changes in the transmission dynamics of infectious disease, potential drivers, geographical distribution, and emergence of novel pathogens. The successful implementation of this response requires not only an evolution from traditional control approaches but also the introduction of novel tools that optimize the allocation of available resources [1].

This article analyzed research conducted in Costa Rica involving mathematical, statistical, and computational techniques, taking into account the country’s specific characteristics, the amount of epidemiological data available, the multiple micro-climates present throughout the country, and different demographic variables. In this context, the use of mathematical and statistical models can serve as platforms to integrate extensive collections of data from different sectors, helping to systematize and integrate experience, as well as expertise from different areas of knowledge and disciplines. This includes techniques that a priori had not been extensively used for these purposes. For instance, analyzing the dynamics of past epidemics, such as the results obtained from the use of classical and Bayesian methods in the study of Zika and chikungunya transmission dynamics, evidenced the need to implement a bottom-up communicative approach at all levels of public health strategies, where more active participation of the community, specifically with individuals that spend most of their time inside their homes, could translate in a greater interest of the population to implement the recommended mosquito control strategies according to the economic, social and epidemiological context of the communities. It also highlighted the importance of continued laboratory-based surveillance in arboviral diseases with similar clinical manifestations.

While the classical and Bayesian methods allowed to comprehend the transmission dynamics of past outbreaks, the use of machine learning algorithms and climate information showed that, by using historical epidemiological and meteorological data, as well as by incorporating lag periods, the Generalized Additive Model and the Random Forest approach cold be used as a tool in the effort to develop early warning system models for Costa Rica and other countries in the region [9]. In this regard, including modeling methods that project tendencies to the information already in use for the decision-making process presents the opportunity to improve the implementation of strategies, the management of programs, and the allocation of available resources. In addition, there is an opportunity for more active inclusion of the population in prevention strategies as sharing information in advance of increased risk during specific periods with members of the community generates the possibility of greater interest to participate in vector control activities.

Therefore, the predictive capabilities of modeling techniques and the greater use of non-traditional information in the development of vector-borne disease control and prevention strategies allows more comprehensive surveillance that involves: the central level in the context of macro decision making such as the allocation of human, technical and administrative resources, at a regional level in the early identification of possible lack of resources, the tailored design of interventions to specific scenarios, as well as, an approach and inclusion of the community in the identification of priority areas of collaboration with the governments and local instances.

Moreover, using the multilayer network model developed as a framework to study Covid-19 tendencies in Costa Rica and extrapolating it with human mobility and different levels of human-vector interaction enables the possibility to design scenarios according to various entomological conditions and mobility levels driven by multiple circumstances such as agriculture activities, tourism, or social and political factors.

Public health authorities must be prepared to adjust to the future challenges that vector-borne diseases may present, both driven by external factors such as climate variability or the emergence of novel pathogens such as SARS-CoV-2, which, health, economic, and social impact has significantly constrained the already limited resources. Hence, a preventive rather than reactive approach should be the basis for the formulation of public health strategies, where modeling techniques serve as cost-effective tools by health authorities.

## Data Availability

All data presented is publicly available.

## Acknowledgements

The authors would like to thank the Research Center in Pure and Applied Mathematics and the School of Mathematics at Universidad de Costa Rica for their support during the preparation of this manuscript.

